# Predicting Seizures Episodes and High-Risk Events in Autism Through Adverse Behavioral Patterns

**DOI:** 10.1101/2024.05.06.24306938

**Authors:** Yashar Kiarashi, Johanna Lantz, Matthew A Reyna, Conor Anderson, Ali Bahrami Rad, Jenny Foster, Tania Villavicencio, Theresa Hamlin, Gari D Clifford

## Abstract

**Objective:** To determine whether historical behavior data can predict the occurrence of high-risk behavioral or seizure events in individuals with profound Autism Spectrum Disorder (ASD), thereby facilitating early intervention and improved support. To our knowledge, this is the first work to integrate the prediction of seizures with behavioral data, highlighting the interplay between adverse behaviors and seizure risk.

**Approach:** We analyzed nine years of behavior and seizure data from 353 individuals with profound ASD. Using a deep learning-based algorithm, we predicted the following day’s occurrence of seizure and three high-risk behavioral events (aggression, self-injurious behavior (SIB), and elopement). We employed permutation-based statistical tests to assess the significance of our predictive performance.

**Main Results:** Our model achieved accuracies 70.5% for seizures, 78.3% for aggression, 80.2% for SIB, and 85.7% for elopement. All results were significant for more than 85% of the population. These findings suggest that high-risk behaviors can serve as early indicators, not only of subsequent challenging behaviors but also of upcoming seizure events.

**Significance:** By demonstrating, for the first time, that behavioral patterns can predict seizures as well as adverse behaviors, this approach expands the clinical utility of predictive modeling in ASD. Early warning systems derived from these predictions can guide timely interventions, enhance inclusion in educational and community settings, and improve quality of life by helping anticipate and mitigate severe behavioral and medical events.

## 1. Introduction

Autism Spectrum Disorder (ASD) is characterized by difficulties in social communication and interactions and the presence of restricted and repetitive behavior patterns Roehr (2013). The prevalance of ASD continues to rise, affecting one in 36 children in the United StatesChristensen et al. (2018). There is a broad spectrum of abilities observed within this population. Many individuals with this diagnosis live a fulfilling independent life, while others require 24-hour support to function and maintain safety. Professional and parent advocates recently began referring the latter group as having “profound” autism. This diversity underscores the importance of a personalized approach to care and intervention, tailored to the unique needs and strengths of each individual with ASD.

### 1.1. High-Risk Behaviors

In addition to diverse neurodevelopmental challenges associated with ASD, challenging behaviors often emerge that interfere with daily functioning Kiarashi et al. (2024); Richler et al. (2006); Buschbacher and Fox (2003); Rad et al. (2025). A range of impact exists from mild disruption to high-risk behaviors that have the potential to cause injury or even death such as aggression, elopement (wandering or bolting away from supervision), pica (ingestion of inedible objects or poisonous fluids), and self-injurious behaviors (SIB).

Self-injurious behaviors have the potential to result in tissue damage, broken bones, and contusions. Concussions and retinal detachment resulting in permanent loss of vision may occur in the case of head-directed SIB. Emerging research into sports where athletes sustain frequent hits to the head shows an association between repeated head trauma and long-term neurological conditions such as chronic traumatic encephalopathy (CTE) Mez et al. (2017). Outcomes of CTE include mood disorders, cognitive decline, memory problems, poor impulse control, and aggression Stern et al. (2011). It is logical that this line of research applies to head-directed SIB as well with an even more detrimental impact when there is an existing ASD disability.

Elopement is another high-risk behavior with potentially tragic outcomes. For individuals who lack safety awareness, elopement has resulted in drowning, death from hyperthermia or hypothermia, and being struck by vehicles or trains McIlwain and Fournier (2017).Restrictive environmental modifications are often needed to ensure safety such as fencing, window locks or blocks, door alarms, and interior bolt-locks. Some people with elopement may require GPS or radio tracking devices.

Aggression may cause injury to both the person engaging in the behavior and those who are intervening to maintain safety. Injuries from aggression result in extended time away from work or permanent disability for workers in an industry already plagued by staffing shortages and frequent turnover. Workman’s compensation claims can be costly for agencies serving those with high-risk behaviors. Aggression often leads to the need for physical interventions that can be traumatizing and harm trust and relationship building with care partners.

While in some cases, the underlying causes of these high-risk behaviors may be difficult to determine, many times they stem from a combination of environmental and internal factors along with underlying skill deficits, particularly in communication and self-regulation. As part of best practices, behavioral clinicians conduct functional behavior assessments to evaluate variables that contribute to a behavior of concern. Despite this technology, it is not always possible to predict whether a particular set of circumstances will trigger a behavior on a given day or if a mild versus a more severe form of a behavior will occur. Understanding and addressing high-risk behaviors is vital, as they significantly impact the quality of life for many individuals with ASD and their care partners.

### 1.2. Seizure

Reflecting the complexity of ASD, individuals with ASD frequently encounter a spectrum of co-morbid medical issues, including sleep disturbances Allik et al. (2008); Anders et al. (2011); Rzepecka et al. (2011); Cohen et al. (2017), sensory sensitivities Talay-Ongan and Wood (2000); Cermak et al. (2010), gastrointestinal disorders Hsiao (2014); Coury et al. (2012), and seizure disorders Volkmar and Nelson (1990); Frye et al. (2016). Co-morbid psychiatric conditions are also common among those with ASDMutluer et al. (2022). These co-morbid medical conditions increase complexity, adding to treatment challenges and significantly affecting the quality of life of those with ASD Mannion and Leader (2013). Among these, seizure disorders represent a particularly complex challenge, standing out for their critical implications on health and well-being compared to other co-morbidities Hirvikoski et al. (2016); Wolpert et al. (2022). The high frequency of seizure episodes in individuals with ASD Hirvikoski et al. (2016) necessitates urgent and effective management strategies. Immediate use of rescue medications is often essential for effective control of these episodes. Given the additional challenges individuals with ASD face, such as communication and behavioral issues, seizures introduce further complexity to their care. There’s a critical need for swift intervention during seizures to mitigate their effects. Enhancing seizure prediction could lead to efficient management, diminishing the impact of seizures on both healthcare systems and the individuals’ well-being, thus highlighting the significance of predictive models and well-structured care plans in improving the management of ASD.

Research highlights a significant correlation between the incidence of seizures and the diagnosis of ASD Kaufmann et al. (2017); Minshawi et al. (2014), particularly among children and adolescents. A hypothesis could be that sensory sensitivities, often observed in individuals with ASD, might serve as predictive indicators for seizure episodes, suggesting a connection where heightened sensory processing challenges precede seizure activity Marco et al. (2011). This insight into the relationship between sensory sensitivities and seizures underscores the critical need for predictive models that can preemptively identify and mitigate these high-risk events.

Additionally, children with ASD are notably more likely to be admitted to the hospital following an emergency department visit for seizure-related disorders, with 4.7% Wolpert et al. (2022)of such visits related to seizure disorders. This statistic not only reflects the severe impact of seizures on this population but also points to the broader implications for healthcare systems and families Thompson and Upton (1992); Kerr et al. (2011). The ability to predict and manage seizure episodes in individuals with ASD could significantly reduce emergency department visits and hospital admissions, thereby improving outcomes.

Various approaches have been explored to predict seizures, with a primary method involving the analysis of electroencephalography (EEG) data to identify patterns or anomalies indicative of upcoming seizures Mirowski et al. (2009); Debicki (2017); Shen et al. (2013); Nasiri and Clifford (2021); Einizade et al. (2023); Regalia et al. (2019); Leijten (2018); Abbasi and Goldenholz (2019); Regalia et al. (2019). Beyond traditional EEG methods, recent advancements in seizure forecasting have leveraged machine learning to enhance algorithmic accuracy and have investigated non-EEG-based indicators, incorporating heart rate variability Jeppesen et al. (2019), in-ear EEG signals Joyner et al. (2024), and electromyography from biceps muscles Beniczky et al. (2018), environmental factors Schelter et al. (2010), and cyclic seizure patterns Karoly et al. (2020); Gleichgerrcht et al. (2022). In addition, stress levels, heart rate variability, and sleep quality have been identified as promising non-invasive markers to monitor seizure susceptibility over extended periods Stirling et al. (2020). Although methods like EEG, fMRI, or sMRI provide high accuracy in controlled settings, they are difficult to scale due to expensive equipment, high costs, and the need for specialized staff.

### 1.3. High-Risk Event Prediction

Despite the promising advancements in models for predicting high-risk medical and behavioral events, several limitations and challenges hinder their application, particularly among children with ASD. Firstly, the continuous and long-term use of EEG devices for seizure detection can be impractical and not well-tolerated, especially for children with the sensory sensitivities common in ASD. These sensitivities may lead to discomfort or distress, making consistent device wear challenging. Secondly, challenging events forecasting aims to estimate the likelihood of an occurrence of the event on any given day, offering a potentially more practical approach over predicting the precise timing of the next seizure.

The necessity for non-invasive and sensory-friendly alternatives is therefore apparent, as traditional EEG devices, with their wires and electrodes, can be particularly bothersome for individuals who are sensitive to tactile sensations. This challenge underscores the need for new approaches that can accommodate the unique needs of individuals with ASD, ensuring that seizure prediction methods are both effective and comfortable for the population. Secondly, available techniques mainly focused on developing prediction algorithms to manage episodic seizures or high-risk behavioral events. Yet, the application of these algorithms in clinical settings has been scarcely examined Freestone et al. (2017); Ferina et al. (2023). In contrast, forecasting aims to estimate the likelihood of a seizure occurring on any given day, offering a potentially more practical approach than predicting the precise timing of the next seizure.

In this work, we introduce a novel model that utilizes the history of challenging behaviors from 353 individuals with profound ASD over a period of nine years to assess the risk of high-risk medical and behavioral events the following day, including seizure episodes, SIB, aggression and elopement. To the best of our knowledge, this is the first work to predict seizure events using historical behavioral data alongside other high-risk behaviors, providing insight into how these behaviors may interact and potentially trigger seizures and other adverse outcomes. By examining the interplay between different challenging behaviors, we aim to offer a more comprehensive and reliable predictive framework, thereby informing earlier interventions and improved support strategies.

## 2. Data Collection

The study was conducted at The Center for Discovery in New York State (TCFD), which provides comprehensive educational, medical, clinical, and residential services to individuals with profound autism and other severe, complex disabilities. All participants required residential care due to the severity of their conditions and their need for intensive support.

We analyzed an existing set of de-identified data routinely collected at TCFD. All included individuals had a previously established ASD diagnosis, often accompanied by intellectual disabilities in the moderate to profound range, limited verbal communication, and significant support needs. The population comprised both children and adults, spanning multiple age groups from pre-adolescence through adulthood, and representing diverse ethnic backgrounds (see Table 1).

**Table 1:** Distribution of participants across life stage and ethnicities by sex. Life stages were defined as Pre-Adolescence (*<* 12 years), Adolescence (12–17 years), Early Adulthood (18–29 years), and Adult (≥ 30 years).

| Category | Age/Ethnicity | Female | Male | Total |
| --- | --- | --- | --- | --- |
| Life stage | Pre-Adolescence | 3 | 19 | 22 |
|  | Adolescence | 19 | 85 | 104 |
|  | Early Adulthood | 36 | 166 | 202 |
|  | Adult | 14 | 33 | 47 |
| Ethnicity | African-American | 7 | 26 | 33 |
|  | Caucasian | 54 | 194 | 248 |
|  | Hispanic | 4 | 22 | 26 |
|  | Other | 6 | 40 | 46 |

The study utilized two datasets. Dataset A consisted of challenging behavior observations continuously collected by trained direct care staff over nine years for 353 individuals. Data were recorded across three shifts per day (morning: 7:00–15:00, afternoon: 15:00–23:00, and overnight: 23:00–7:00) and included a range of behaviors such as Aggression, Disruptive Behavior, Elopement, Self-Injurious Behavior (SIB), Impulsive Behavior, Agitation, Mouthing/Pica, Property Destruction, Task-Refusal, Inappropriate Touch, and restricted/repetitive behaviors.

Dataset B included seizure episode durations and subsequent recovery times for 55 individuals, recorded during daytime hours. Both datasets were derived from the same residential population at TCFD, with all participants in the Dataset B also included in the Dataset A. Table1 shows the demographic information for Dataset A (inclusive dataset). This study was approved by the TCFD and Emory Institutional Review Boards (STUDY00003823: ‘Predicting Adverse Behaviour in Autism’).

## 3. Methods

### 3.1. Preprocessing

In the preprocessing stage for the challenging behavior dataset, we began by identifying the top 7 most prevalent behaviors across our entire study population as indicated above. Any behaviors not fitting these categories were grouped under the label Other, resulting in a framework with 8 distinct behavior types for each recorded episode (i.e., an event that occurred in the morning, afternoon, or evening shifts). By aggregating the labels across different times of the day, we generated a binary vector with 8 entries for each day for every participant. Given our focus on predicting aggression, SIB and elopement as high-risk behavioral events, we excluded records from individuals without any incidents of agression, SIB or elopement. Consequently, our refined dataset included records from 277 individuals with Aggression, 192 with SIB and 125 with elopement. For the seizure dataset, we initially identified individuals featured in both datasets (the population of this dataset is subset of the first dataset). Subsequently, we introduced a binary feature indicating the presence or absence of a seizure episode for a given individual on a specific day, resulting in a binary vector with 9 entries for each day (see Fig. 1. This process yielded a dataset that included 55 individuals.

**Figure 1:**
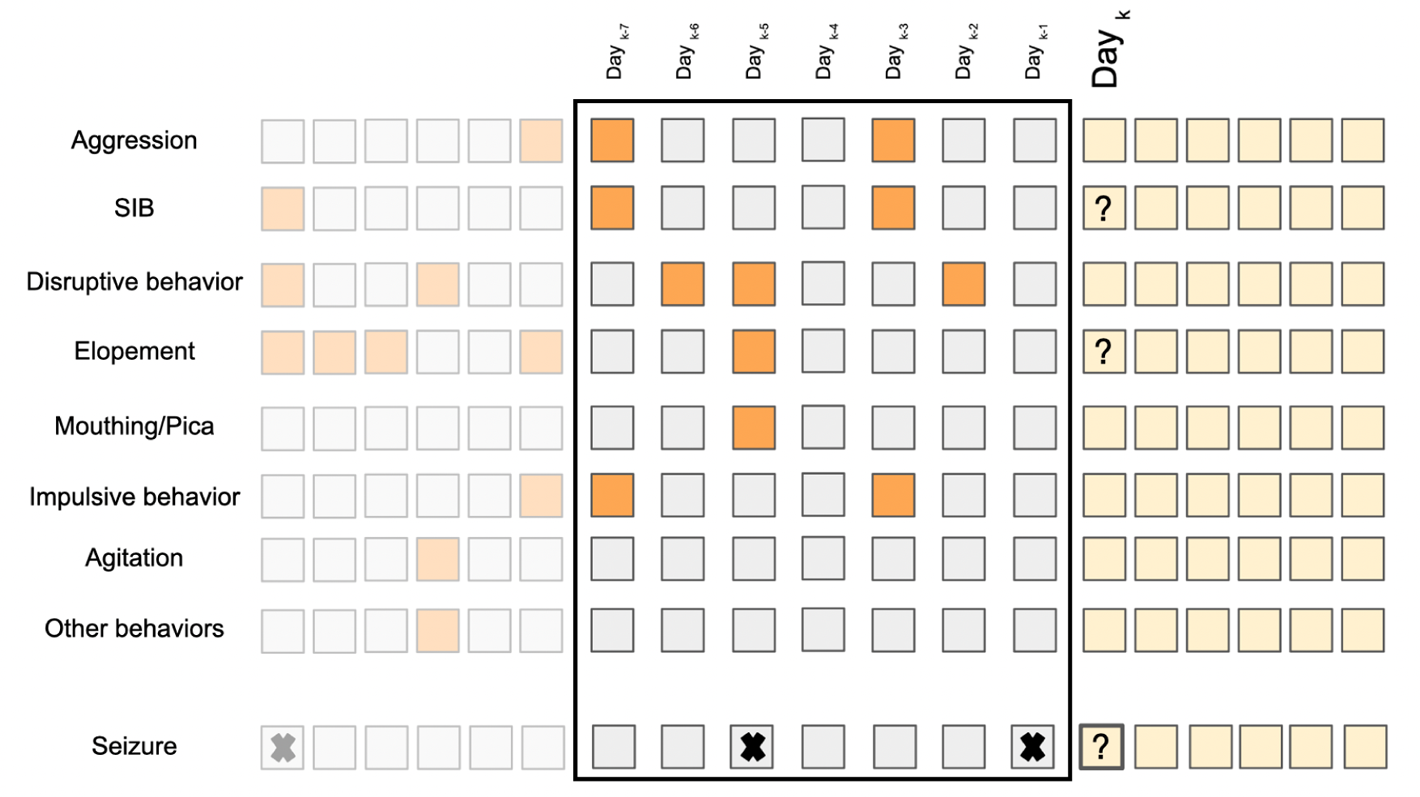
The feature vector is composed of seven selected behaviors, determined to be most prevalent across the study population, along with any other observed behaviors, each represented in binary form. This information forms a two-dimensional feature vector covering a timespan of the past seven/fourteen days, with the label indicating the occurrence of challenging behaviors and seizure episode on the subsequent day. Cells with lower opacity represent records from previous days, the feature vector consists of the cells enclosed by the black rectangle, and the yellow cells represent the upcoming days for which we aim to predict the presence of high-risk events.

In forming the input features for each participant, as illustrated in Fig. 1, we used two time windows. The first approach involved using data from the 7 days leading up to an event to forecast aggression/SIB/elopement/seizure occurrences on the following day. The second approach extended this time window by using data from the previous 14 days for the same task.

### 3.2. Prediction Model

In this paper we used a CNN architecture with two-dimensional convolutional layers to extract spatial features effectively. It integrates batch normalization layers to maintain stable learning conditions and employs max pooling layers to decrease data dimensionality. The architecture is further enhanced with dense layers activated by the ReLU function, which are instrumental in identifying nonlinear relationships. To mitigate the risk of overfitting, a dropout layer is incorporated. The architecture that was designed for predicting the likelihood of presence or absence of each category of event is depicted in Fig. 2.

**Figure 2:**
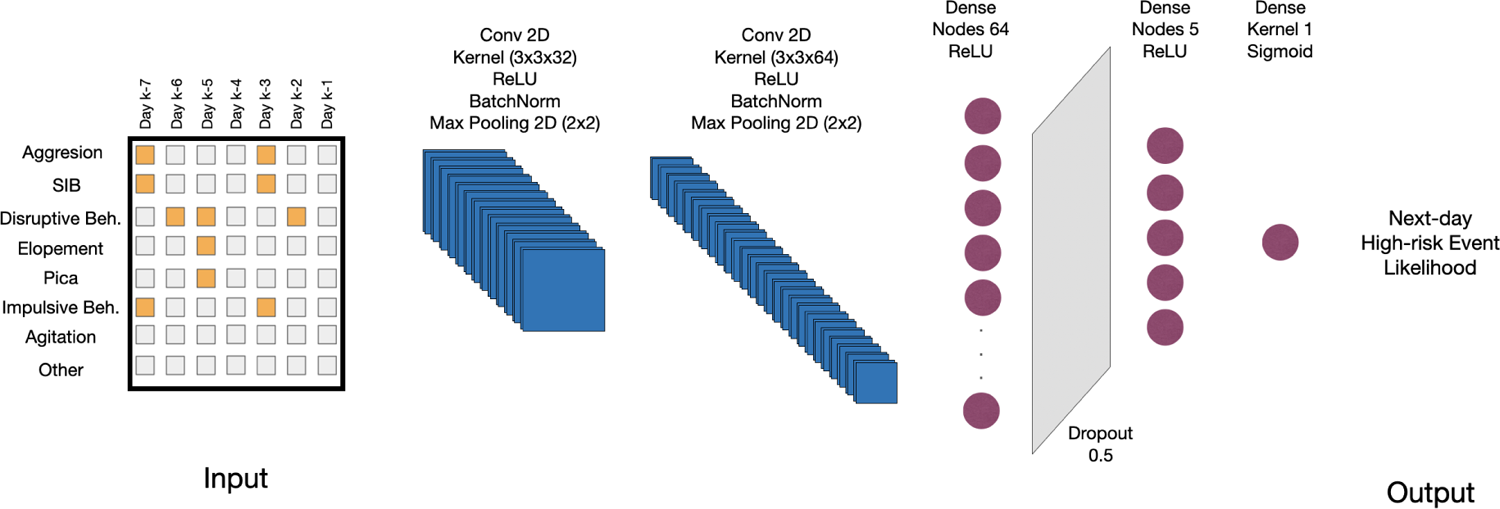
The diagram illustrates the deep learning model designed for the binary prediction of the occurrence of high-risk behavioral and medical events on the following day. The model architecture includes two-dimensional convolutional layers, batch normalization, max pooling, dense layers with ReLU activation, a dropout layer for regularization, and a final dense layer with a sigmoid activation function for outputting the likelihood for an individual displaying a given behavior (e.g., self-injurious behavior, elopement, or seizure episode) the following day.

In this paper, we used a CNN architecture composed of two-dimensional convolutional layers and associated operations to effectively extract spatial features. Specifically, the first convolutional layer employs 32 filters with a kernel size of 3 × 3 and ReLU activation, followed by batch normalization to stabilize training and max pooling (2 × 2) to reduce spatial dimensions. The second convolutional layer uses 64 filters with a kernel size of 2 × 2 and ReLU activation, again followed by batch normalization and max pooling (2 × 2). After these convolutional and pooling operations, the feature maps are flattened and passed through a dense layer with 64 units and ReLU activation. To mitigate the risk of overfitting, a dropout layer with a rate of 0.5 is incorporated. Finally, the model culminates in a single-unit dense layer activated by a sigmoid function to predict the likelihood of events. The model is trained for 50 epochs using the Adam optimizer with a batch size of 32 and binary cross-entropy loss. A schematic representation of the architecture, detailing layer dimensions, kernel sizes, activation functions, and other hyperparameters, is presented in Fig. 2.

### 3.3. Evaluation Metrics

The training procedure for these models involved allocating 80% of the data for training, while the remaining 20% was set aside for testing purposes. This approach, including both training and evaluation, was conducted in a subject-specific manner, aiming to preserve the temporal causality while assessing the predicting scores through a individual-specific analysis.

To evaluate the effectiveness of our model, we utilized area under the receiver operating characteristic curve (AUROC), area under the precision-recall curve (AUPRC), accuracy, and *F*_1_ score. These metrics were computed through macro-averaging across all subjects and evaluations are presented as mean ± standard deviation for individuals. Basically, we computed the metrics for each individual and found the population mean and standard deviation across the population of individuals. For assessing the statistical significance of the model’s accuracy over randomness, we chose accuracy as the primary metric due to its relevance to the specific classification tasks. We employed permutation testing as detailed in Algorithm 1. This method shuffles the labels of the test set to generate distributions of accuracy under the null hypothesis that our model’s performance is comparable to random guessing (i.e., prevalence aware guessing). The performance of our model is deemed statistically significant if it surpasses the accuracy benchmarks for a specific individual, as determined by comparing the actual model’s performance against this distributions.

In order to assess whether the model’s performance exceeds what could be expected from a baseline that relies solely on previous nights’ behavioral prevalence, we employed a permutation-based statistical test. This approach preserves the underlying prevalence of behaviors, reflecting a scenario in which predictions are derived exclusively from historical frequencies rather than from model-driven feature extraction. Specifically, it mirrors how a staff member might estimate the likelihood of a behavior by referencing its past frequency, without leveraging current context or additional signals.

**Algorithm 1.**
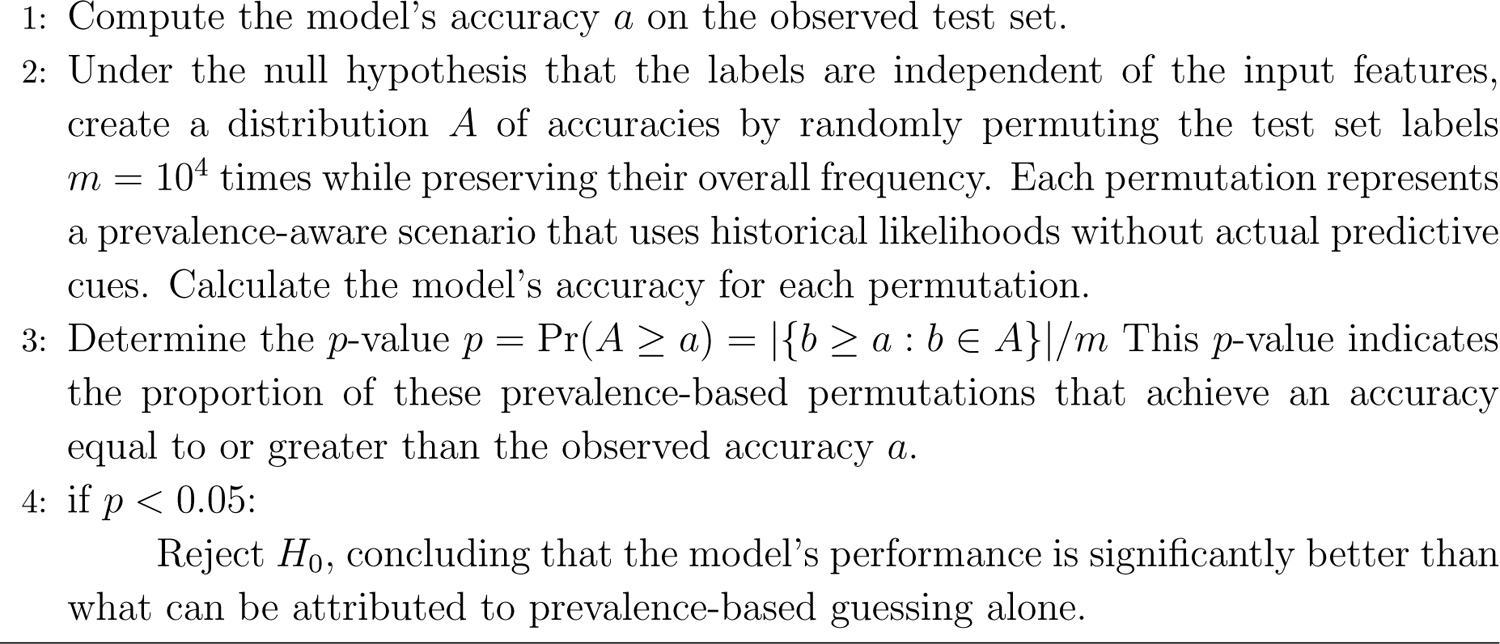
Statistical Test.

To further quantify the model’s improvement over this prevalence-aware baseline, we introduce the ΔAccuracy, defined as the average increase in accuracy relative to the expected accuracy under the null hypothesis. This metric serves as an effect size, highlighting the practical significance of the model’s predictive ability beyond what could be achieved by relying solely on historical frequencies.

*Feature importance* Here we explore the feature importance in our CNN model, designed for the binary prediction of high-risk behavioral and medical events the following day. We utilized Gradient-weighted Class Activation Mapping (Grad-CAM) Selvaraju et al. (2017) on the first convolutional layer of the CNN. Contrary to traditional applications that target deeper layers, focusing on the first layer allowed us to understand the initial feature extraction process directly related to the input data. Grad-CAM generates a coarse localization map, visually highlighting the significant regions in the input image that influence the model’s prediction. The method is formulated as follows:

i. Compute the gradient of the class score, *Y^c^* (here did the analysis for positive class *c* = 1), with respect to the feature maps, *A^k^* (*k* here is the index of the filter), of the first convolutional layer to obtain 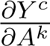.
ii. Calculate the neuron importance weights, *α^c^*, through average pooling of these gradients, expressed as 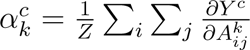, where *Z* represents the total number of pixels (i.e., features) in a feature map, and *i, j* are the pixel indices.
iii. Produce the Grad-CAM heatmap by applying a weighted combination of these activation maps and a ReLU function: 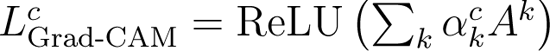.

## 4. Results

### 4.1. Performance Analysis

The confusion matrices presented in Figure 3 illustrate the model’s predictive micro performance across a cohort of individuals exhibiting high-risk behaviors. Specifically, Seizure events, were predicted with accuracies of 70.6% and 71.6% for 7-day and 14-day. For aggression the model demonstrated micro accuracies of 80.9% and 84.8% using historical behaviors 7-day and 14-day time windows, respectively, with corresponding micro F1 scores of 0.74 and 0.79. For SIB, the model achieved micro accuracies of 82.8% and 85.4%, and micro F1 scores of 0.75 and 0.79 for the same windows sizes. In predicting elopement, the model reached its highest micro accuracies of 85.7% and 88.9% for the 7-day and 14-day, respectively, accompanied by micro F1 scores of 0.76 and 0.81. These results indicate that utilizing 14 days of historical data leads to slightly higher accuracy and F1 scores across all tasks.

**Figure 3:**
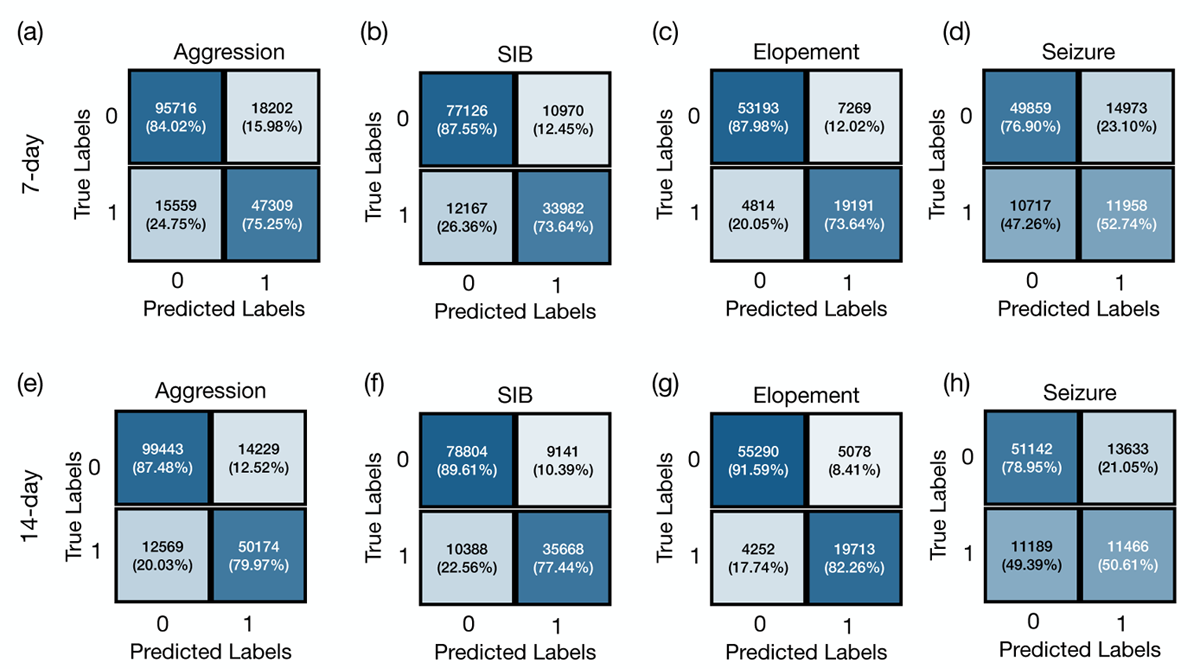
Confusion matrices for predicting aggression, self-injurious behavior (SIB), elopement, and seizure. Sub-tables (a), (b), (c), and (d) illustrate confusion matrices predict these behaviors based on a 7-day historical period. Sub-tables (e), (f),(g), and (h) predict these behaviors based on a 14-day period.

Table 2 presents the mean and standard deviation (SD) of the macro F1 scores and macro accuracies for each event across different time frames. For seizure episode predictions the model achieved an F1 score of 0.77 across both time windows and a slight improvement in accuracy from 69.2% ± 7.3% to 70.5% ± 6.5%. For aggression, the F1 score was 0.77 ± 0.20 over 7 days, which slightly increased to 0.80 ± 0.20 over 14 days. The model’s accuracy for predicting aggression also rose from 75.8% ± 14.7% to 78.3% ± 16.4% as the prediction window extended. In the case of SIB, the F1 scores were 0.82 ± 0.15 for the 7-day and 0.83 ± 0.19 for the 14-day predictions, with accuracies of 78.7% ± 14.2% and 80.2% ± 17.5%, respectively. For elopement, there was an increase in the F1 score from 0.85 ± 0.18 at 7 days to 0.89 ± 0.13 SD at 14 days, and accuracy improved from 81.8% ± 15.7% to 85.7% ± 11.2%.

**Table 2:** Performance metrics for predicting next-day high-risk behavior and seizure using historical records of the previous 7 and 14 days (*Mean* ± *SD*). Significance Achieved represents the number of cases in the population for which we could reject the null hypothesis. ΔAccuracy indicates the margin by which our model outperforms the baseline.

| Event | Window | F<sub>1</sub> Score | Accuracy (%) | Significance | ΔAccuracy (%) |
| --- | --- | --- | --- | --- | --- |
| Seizure | 7 days | $0.77 \pm 0.14$ | $69.2 \pm 7.3$ | 54 from 55 | $10.5 \pm 4.7$ |
| Seizure | 14 days | $0.77 \pm 0.10$ | $70.5 \pm 6.5$ | 51 from 55 | $10.3 \pm 5.3$ |
| Aggression | 7 days | $0.77 \pm 0.20$ | $75.8 \pm 14.7$ | 236 from 277 | $12.8 \pm 10.3$ |
| Aggression | 14 days | $0.80 \pm 0.20$ | $78.3 \pm 16.4$ | 234 from 276 | $14.7 \pm 11.6$ |
| SIB | 7 days | $0.82 \pm 0.15$ | $78.7 \pm 14.2$ | 168 from 192 | $14.3 \pm 11.1$ |
| SIB | 14 days | $0.83 \pm 0.19$ | $80.2 \pm 17.5$ | 168 from 192 | $15.5 \pm 12.5$ |
| Elopement | 7 days | $0.85 \pm 0.18$ | $81.8 \pm 15.7$ | 96 from 125 | $15.7 \pm 12.4$ |
| Elopement | 14 days | $0.89 \pm 0.13$ | $85.7 \pm 11.2$ | 96 from 124 | $17.5 \pm 13$ |
| Severe Aggression | 7 days | $0.79 \pm 0.13$ | $70.0 \pm 15.1$ | 3 from 7 | $2.3 \pm 1.2$ |
| Severe Aggression | 14 days | $0.83 \pm 0.09$ | $73.8 \pm 13.3$ | 3 from 7 | $0.7 \pm 1.3$ |
| Severe SIB | 7 days | $0.78 \pm 0.09$ | $68.4 \pm 10.2$ | 2 from 5 | $4.1 \pm 2.2$ |
| Severe SIB | 14 days | $0.82 \pm 0.09$ | $74.0 \pm 11.4$ | 3 from 5 | $3.25 \pm 1.8$ |

To investigate the potential difference in model effectiveness across sexes, we analyzed the performance metrics for four key events (Seizure, Aggression, SIB, and Elopement using F_1_ scores and accuracy as evaluation criteria. As shown in Table 3, the results demonstrate minimal differences in performance between female and male participants. For instance, the F_1_ scores for Seizure detection are 0.72 ± 0.10 for females and 0.78 ± 0.10 for males, while accuracy values are 70.0% ± 7.6% and 70.6% ± 6.1%, respectively. Similarly, in Aggression detection, the F_1_ scores and accuracy metrics for both sex show overlapping standard deviations, with no substantial disparity.

**Table 3:** Performance metrics (*Mean* ± *SD*) for different events using historical records of the previous 14 days across sexes.

| Event | Sex | F<sub>1</sub> Score | Accuracy (%) | Significance | ΔAccuracy (%) |
| --- | --- | --- | --- | --- | --- |
| Seizure | Female | $0.72 \pm 0.10$ | $70.0 \pm 7.6$ | 7 from 8 | $11.1 \pm 4.7$ |
| | Male | $0.78 \pm 0.10$ | $70.6 \pm 6.1$ | 44 from 47 | $10.2 \pm 5.4$ |
| Aggression | Female | $0.77 \pm 0.25$ | $76.7 \pm 19.2$ | 41 from 50 | $14.3 \pm 11.3$ |
| | Male | $0.81 \pm 0.18$ | $78.7 \pm 15.8$ | 190 from 226 | $14.8 \pm 11.6$ |
| SIB | Female | $0.83 \pm 0.20$ | $80.6 \pm 17.6$ | 36 from 41 | $15.1 \pm 11.5$ |
| | Male | $0.83 \pm 0.19$ | $80.1 \pm 17.5$ | 132 from 151 | $15.6 \pm 12.8$ |
| Elopement | Female | $0.89 \pm 0.10$ | $84.7 \pm 11.6$ | 9 from 13 | $18.7 \pm 12.6$ |
| | Male | $0.89 \pm 0.13$ | $85.9 \pm 11.2$ | 87 from 111 | $17.6 \pm 13.3$ |

### 4.2. Statistical Analysis

We applied the significance test described in subsection 3.3 to assess whether the model’s performance for each individual was statistically significant. The number of instances where the null hypothesis (stating that the model performs no better than permuted labels) was rejected is reported. For in 54 out of 55 cases for the 7-day period and in 51 out of 55 cases for the 14-day period. aggression, significance was achieved in 236 out of 277 cases over a 7-day window and 234 out of 276 cases over a 14-day window. For SIB, the counts were 168 from 192 for both timeframes. For elopement, the model’s predictions were significant in 96 out of 125 cases for the 7-day period and 96 out of 124 cases for the 14-day period. Finally, for seizure predictions, the null hypothesis was rejected. The marginal improvements in ΔAccuracy across behaviors suggest that extending the historical data window to 14 days slightly enhances predictive performance, though the magnitude of these effects varies.

Table 2 also details results for predicting *severe* aggression and *severe* SIB in a small cohort, where at least 10% of the target behavior was recorded as severe. For severe aggression, the model achieved an F1 score of 0.79 ± 0.13 with an accuracy of 70.0%±15.1% over a 7-day window, which increased slightly to an F1 score of 0.83±0.09 and an accuracy of 73.8% ± 13.3% over 14 days. Predictions for severe SIB followed a similar trend, with F1 scores rising from 0.78 ± 0.09 to 0.82 ± 0.09 and accuracy increasing from 68.4% ± 10.2% to 74.0% ± 11.4%. However, the effect sizes, as measured by ΔAccuracy, were small, ranging from 2.3% ± 1.2% to 3.3% ± 1.8%. Significance was achieved in only a few cases for severe aggression and severe SIB, highlighting the limited success of the model in predicting these extreme behaviors.

Table 3 shows that the model’s performance is consistent across sexes, with similar effect sizes and significance achieved for both males and females. In predicting Seizure, the ΔAccuracy is nearly identical for females (11.1% ± 4.7%) and males (10.2% ± 5.4%), and a similar pattern is seen for SIB and elopement. The number of cases where significance was achieved is also comparable across sexes, indicating no significant disparities. Overall, these results suggest that the model performs equally well for both sexes, ensuring fairness and reliability in its predictions.

### 4.3. Feature Importance Analysis

Figure 4 presents the impact of each feature using Grad-CAM, presented in the methods section (see 3.3); we observed that disruptive behavior plays a crucial role in predicting all high-risk events, underscoring its significance across different high-risk behaviors. Disruptive behaviors represent a broad category inclusive of less impactful behaviors such as screaming, dropping, hitting or throwing objects, or disrobing, to provide a few examples. Our findings suggest that there is a pattern of lower impact behaviors that precede aggression, self-injury, elopement, and seizures.

**Figure 4:**
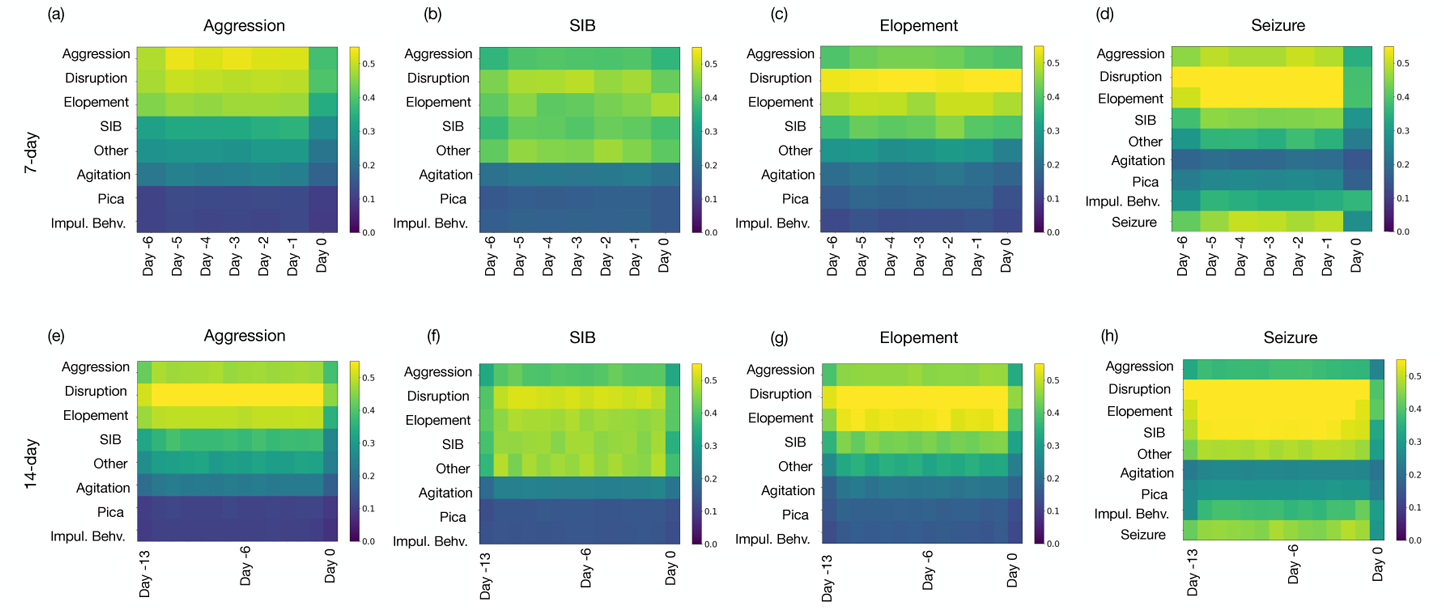
Representation of features importance using GradCAM for predicting high-risk events. Day −*j* refers to the *j^th^* day preceding the target day for which the prediction is being made.(a), (b), (c), and (d) illustrate the feature importance rankings for predicting aggression, SIB, elopement, and seizures, respectively, based on a 7-day window. (e), (f), (g), and (h) extend the analysis to a 14-day window for the same outcomes. The comparison highlights how the predictive value of specific features shifts with the extension of the historical data period.

**Figure 5:**
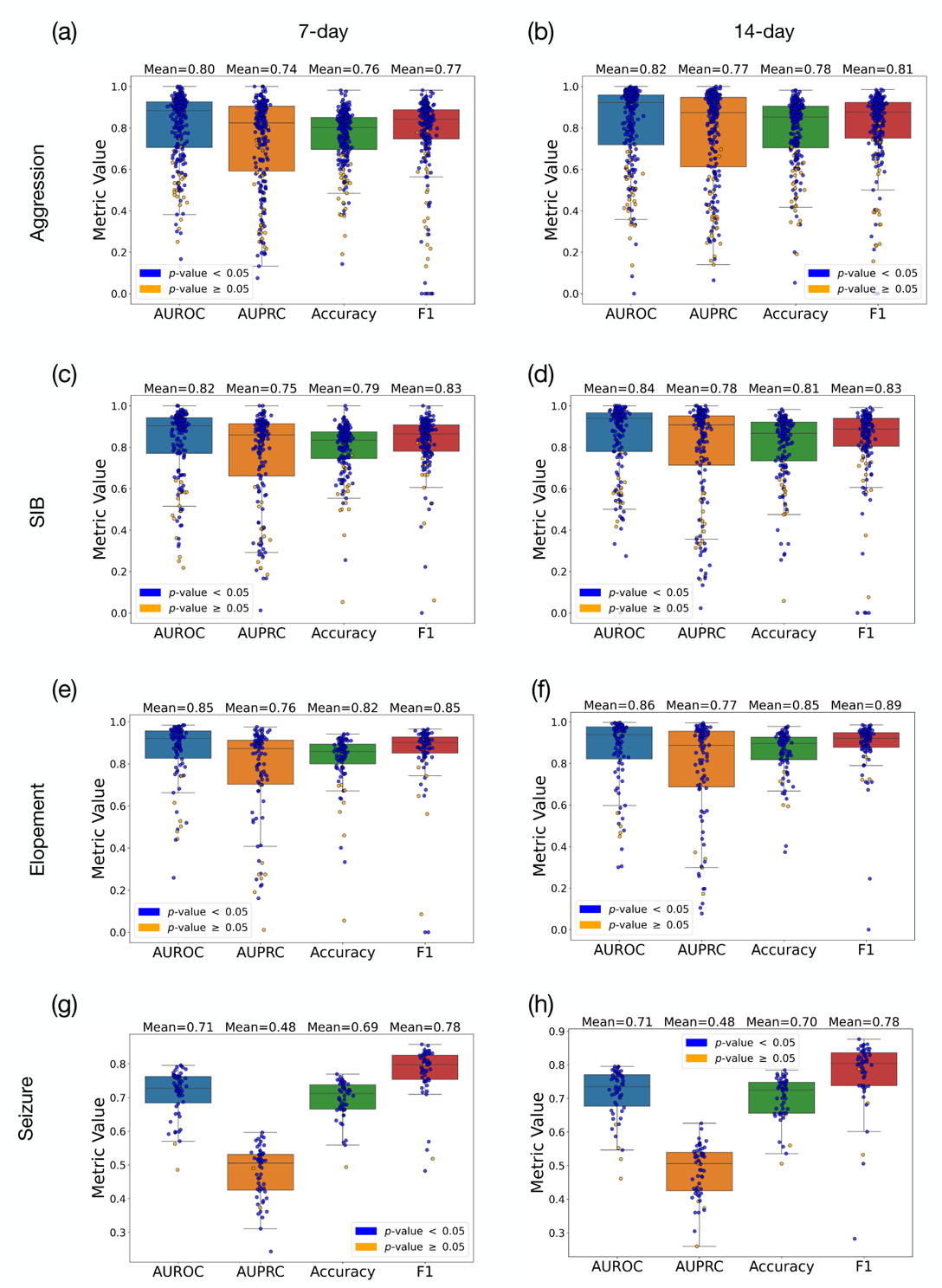
Representation of prediction results using 7-day and 14-day spans of prior historical data for predicting SIB, elopement, and seizure, presenting AUROC, AUPRC, accuracy, and F1 score. Blue and yellow circles represent cases where we achieved, and could not achieve, statistical significance, respectively. Subfigures (a), (c), (e), and (g) depict the results for aggresion, SIB, elopement, and seizure using 7-day data, while (b), (d), (f), and (h) illustrate the results for SIB, elopement, and seizure using 14-day data.

Behaviors that occur together and result in similar consequences are referred to as belonging to the same response class. Previous research has demonstrated associations between low- and high-risk behaviors such as restricted and repetitive behaviors and aggression Gohari et al. (2024). To a lesser extent, our study reveals that the presence of one high-risk behavior can predict another high-risk behavior. The co-occurrence of high-risk behaviors including self-injury and aggression is well-documented in the literature Matson et al. (2008); Kanne and Mazurek (2010).

Particularly noteworthy is the predictive value that the history of seizure events holds when forecasting future seizures; however, it is not the sole contributor. Our analysis indicates that low-risk disruptive behaviors and high-risk behaviors of elopement, aggression and self-injurious behavior, also exhibit a prominent predictive value in seizure forecasts. Previous research has demonstrated that those with ASD and seizures are more hyperactive and irritable compared to individuals with ASD who do not have seizures Viscidi et al. (2013). In this research, hyperactivity and irritability were operationalized to contain similar behaviors to those defined in our study.

While previous studies have found associations between different types of behaviors and behaviors and seizures, our study adds to the literature in that we demonstrate that the timing and patterns of behaviors over time serve as important predictors of upcoming high-risk behavioral and medical events. Simply knowing the co-occurrence of events is not as clinically actionable as individualized predictions based on what behaviors occur and when. Our study suggests that a multifaceted approach could enhance the accuracy of predicting seizures and other high-risk events.

In terms of understanding more severe forms of behaviors, our results indicate that, since such behaviors are inherently sparse, the models’ ability to predict more severe high-risk behaviors is constrained. This limitation is particularly pronounced due to the lack of additional, pertinent information regarding individual subjects, such as major life events, medication usage, and comorbidities. Therefore, the practical utility of the models for predicting more severe high-risk behavior without incorporating more comprehensive information or features is limited. It underscores the necessity for a more holistic approach in model development that integrates broader aspects of individual profiles to enhance predictive accuracy.

Another observation was as we extended the historical window to 14 days, illustrated in panels (e) through (h), there was a notable shift in the importance of these features. For instance, aggression remained a dominant feature for predicting SIB over both time frames, whereas disruptive behavior was prominent in the 14 days for predicting elopement. This analysis highlights how extending the historical data window can recalibrate the predictive value of specific features, which may refine our predictive models’ accuracy for severe high-risk behaviors.

## 5. Discussion

The behavioral and medical complexity of individuals with profound autism, particularly those prone to seizures, presents unique challenges for care providers. While functional behavior assessments may identify antecedents to high-risk behaviors, these assessments often fail to reliably anticipate events such as seizures, which can occur unpredictably and with severe consequences. Accurate seizure prediction is particularly critical as it allows care partners to take timely measures to reduce the risk of harm.

This paper demonstrates an AI-driven model capable of predicting seizures and high-risk behaviors. As shown in Table 2 and 3. This improvement highlights the model’s potential to deliver actionable insights for real-time interventions, which can significantly reduce the impact of seizure episodes. Integrating this technology into monitoring systems can aid in ongoing risk assessment, allowing healthcare professionals to preemptively address seizure risks and tailor treatment plans to the dynamic nature of epilepsy.

Table 2 shows that using 14 days of historical data leads to consistently better predictive performance compared to 7 days, with noticeable improvements across most behaviors. While the gains are more pronounced for some behaviors like aggression and elopement, even smaller improvements, such as for seizure prediction, align with this overall trend. Current results suggest 7-day time-window is sufficient to form a reliable predictor of the next day high-risk event. Additionally, Table 3 highlights that these improvements are consistent across sexes (for 14 days time window), with no significant disparities observed. This suggests that extending the historical data window enhances accuracy without introducing sex-based biases.

Our findings further emphasize the potential of combining behavioral and physiological data for seizure forecasting. Recent evidence from a comprehensive Canadian survey of 196 patients and 150 caregivers supports the inclusion of behavioral cues in seizure prediction. In the survey, approximately 12% of participants reported the ability to anticipate seizures based on preictal symptoms such as mood changes, dizziness, and cognitive disturbances, sometimes up to 24 hours in advance Larivìere et al. (2020). This underscores the value of integrating non-physiological indicators into predictive models, offering caregivers additional time for preventive measures.

Additionally, the influence of historical behaviors on the prediction of high-risk events, including seizures, was evident in this study. As illustrated in Figure 4, cyclic patterns were observed, where prior occurrences of a specific behavior or medical event, such as a seizure, significantly contributed to predicting future instances. Disruptive behaviors, in particular, were found to have a broad impact on predicting various high-risk events, suggesting they may serve as a general marker of underlying instability. These findings highlight the interconnected nature of behavioral dynamics and the importance of holistic approaches in predictive modeling.

By focusing on seizure prediction, this study not only advances the understanding of preictal patterns but also demonstrates the potential of AI-driven models to transform care for individuals at risk. Deploying and evaluating these models in real-world settings like TCFD offers an opportunity to validate their utility and further refine intervention strategies. The ability to anticipate and mitigate the impact of seizures can improve safety and quality of life for individuals with profound autism, underscoring the importance of integrating predictive technologies into routine care.

## 6. Conclusion

This study demonstrated the feasibility of using a deep learning-based algorithm to predict high-risk behaviors and seizure episodes in individuals with ASD. By analyzing nine years of behavioral and seizure data from 353 individuals, we showed that the history of high-risk events contains valuable information for predicting both next-day behaviors and seizure episodes. Notably, the model demonstrated accuracies of 70.5% for seizures, 78.3% for aggression, 80.2% for SIB, and 85.7% for elopement, with statistical significance achieved for over 85% of the population across all event types. These results highlight the interplay between adverse behaviors and seizure risks, offering new insights into how behavioral patterns can serve as early indicators of seizure episodes.

Our study demonstrates significant advancements over prior methodologies, particularly in comparison to the work by Ferina et al. (2023), which was limited to predicting each behavior using data solely from that specific behavior (e.g., using SIB events to predict SIB). While their model achieved statistical significance for 15-20% of participants, our approach, which leverages the interplay between different behaviors to predict the presence of each adverse behavior, outperforms theirs, achieving statistical significance for over 85% of participants across all behaviors. We believe this improvement stems from our model’s ability to capture the dynamic relationships between behaviors, highlighting the predictive value of using one behavior to infer another. This dynamic interplay represents an important advancement, suggesting that our approach more effectively explains the mechanisms underlying adverse behaviors and their interdependencies, thereby contributing to the existing body of literature.

Our findings also emphasize that a 7-day historical window provides sufficient information to predict next-day high-risk behaviors. The results suggest that while extending the data window to 14 days can enhance accuracy slightly, shorter windows still capture enough behavioral patterns for reliable predictions. This makes the model both practical and efficient for real-world applications. Furthermore, the analysis revealed no significant disparities in predictive performance across sexes, underscoring the fairness and generalizability of the algorithm.

The ability to predict seizure episodes and high-risk behaviors has significant implications for care strategies. For seizures, early predictions allow for proactive measures such as close monitoring, minimizing physical demands, and adapting care plans to reduce risks and potential injuries. For high-risk behaviors, predictions enable environmental and staffing adjustments, implementation of preventive interventions, and mitigation of their impact on both individuals and caregivers. These findings pave the way for AI-driven early warning systems that can transform the care paradigm in ASD, shifting from reactive to anticipatory approaches and improving quality of life for individuals and their care teams.

## Data Availability

The used for this study is available upon reasonable request.

## Acknowledgments

This work was supported by the Center for Discovery. YK, PS, AR and GC were partially supported by James M. Cox Foundation and Cox Enterprises, Inc., in support of Emory’s Brain Health Center and Georgia Institute of Technology. GC is partially supported by the National Center for Advancing Translational Sciences of the National Institutes of Health under Award Number UL1TR002378. YK is partially supported by Thrasher Research Fund Early Career Award Program. The content is solely the responsibility of the authors and does not necessarily represent the official views of the National Institutes of Health, the Center for Discovery, Thrasher Research Fund, or the Cox Foundation.

